# Higher knee hypertension in patients with hemiplegia correlates lower lateral and medial meniscus volume in the paretic leg

**DOI:** 10.1101/2022.02.18.22271154

**Authors:** Wenshan Li, Tiancong Li, Xiaoshuang Xi, Rong Zhang, Weishuang Sun, Conglin Han, Dan Zhang, Weijun Gong

## Abstract

Stroke is a disease with high morbidity and disability rate. After stroke, slower walking velocity, shorter step length, higher cadence, larger step width, and longer stance phase duration can be apparently observed. Additionally, these abnormal gait parameters eventually lead to knee hyperextension, known as genu recurvatum. Walking with such knee hyperextension usually induces knee pain and joint lesions, leading to cumulative damage and degenerative changes and further decreasing the standing phase’s stability. However, do higher angles of knee hyperextension have any correlation between the gait parameters, the kinematics, the kinetic parameters, the meniscus volume, and the water content of meniscus in paretic and non-paretic legs? This study attempted to answer these questions. The results revealed that 1) longer time since hemiplegia led to larger angles of knee hyperextension, 2) larger angles of knee hyperextension led to more tears in medial/lateral meniscus, and 3) larger angles of knee hyperextension decreased water content of the lateral meniscus in the non-paretic leg but increased water content of the medial meniscus. This was the first study to demonstrate these three above-mentioned observations through magnetic resonance imaging and motion capture system. These results suggested that 1) the knee hyperextension needed to be diagnosed and treated as early as possible; 2) the time since hemiplegia could be an indicator of sign of knee hyperextension; and 3) it might be not effective to identify the knee hyperextension by using spatial-temporal gait parameters.

## INTRODUCTION

Stroke is a disease with high morbidity and disability rate, in which about 70% of patients have hemiplegia [1]. Approximately 50∼80% of hemiplegic survivors may regain the walking capability; however, their gait patterns are quite different than the one before suffering hemiplegia [2]. This abnormal gait pattern is characterized by alternations in spatial-temporal gait parameters, such as slower walking velocity, shorter step length, higher cadence, larger step width, and longer stance phase duration [3-4]. Specifically, muscle activations result in gait characteristics in these hemiplegic survivors, resulting in knee hyperextension, known as genu recurvatum [5] during the stance phase. More precisely speaking, among all stroke survivors, 40% ∼ 70% of these patients with hemiplegia suffer from genu recurvatum [6]. This locomotor behavior induces a high knee extensor moment to prevent collapse during the stance phase [6].

Long-term knee hyperextension causes the abnormal load-bearing response of the knee joint. Walking with such knee hyperextension usually induces knee pain and joint lesions, leading to cumulative damage and degenerative changes [7] and further decreasing the standing phase’s stability. The noticeable kinematic gait change due to the hyperextension is the higher peak extensor torque compared to age-matched controls [8]. This higher peak extensor torque is speculated by the weakness of the quadriceps [8]. Importantly, these higher torque values might be the clinical concern because an increase in the extensor moment at the knee likely increases the potential risk for damages to the posterior side of the menisci [9].

For the knee joint, the meniscus plays a critical role in increasing the contact area between the femur and the tibia to enhance the capability of the knee to bear the loading up five to10 times more than its body weight [10]. Specifically, a study uses magnetic resonance technology to show meniscal movement and height changes during knee flexion/extension. During knee extension, the medial and lateral menisci move forward, the range of motion is larger in the lateral meniscus than in the medial meniscus; moreover, the range of motion is larger in the anterior horn than in the posterior horn [10]. This design gradually increases the tibiofemoral contact area to transfer the load uniformly to maintain the stability of knee joint [10]. However, these magnetic resonance studies majorly focus on the healthy controls but not on the hemiplegic survivors. The knowledge of how knee hyperextension affects the meniscus is limited. To understand the correlation between knee hyperextension and volume of meniscus may help physical therapies to develop an advanced rehabilitation protocol.

Moreover, about 50% of stroke patients have proprioception deficiency [11], which comes into interaction inhibition, changing the knee joint stability and causing poor activity control. This proprioception deficiency is highly correlated to the damage of nerve fibers that are innervated to the peripheral portion of menisci and the anterior and posterior horns [12]. In addition, during the knee hyperextension, the meniscal horns are compressed and stressed. These compressions and stress might potentially place the meniscus at a greater risk of injury [12] and further tear the proprioception, the perception of joint motion, and position.

From abovementioned studies, the knee hyperextension led to the impairments in knee joint in these stroke patients had been well-investigated; however, do higher angles of knee hyperextension have any correlation between the gait parameters, the kinematics, the kinetic parameters, the meniscus volume, and the water content of meniscus in paretic and non-paretic legs. This study hypothesized that there was a negative correlation between the angles of knee hypertension and step length, cadence, stance phase, joint angles, ground reaction forces, but there was a positive correlation between the angles of knee hypertension and knee torques. Additionally, this study hypothesized that there was a negative correlation between the angles of knee hypertension and meniscus volume, and the water content of meniscus.

## METHOD

### Ethics Statement

Stroke patients in this study were recruited from Beijing Rehabilitation Hospital of Capital Medical University. This study was approved by Beijing Rehabilitation Hospital of Capital Medical University Ethics committee. (#2021bkkyLW001)

### Participants

Eight patients with chronic hemiplegic (6 male, 2 female) volunteered to participate in this study (age: 50.25 (12.09) years; height: 168.87 (7.05) cm, mass: 70.62 (6.28) kg). Participants were included as follows: participants needed to be over 30 years old, had a hemiplegia following a stroke occurring more than 6 months, had an ability to walk 10 meters without aids, had a Function Ambulation Category level at least 3 and above, had a hemiplegic lower extremity identified as Brunnstrom state III or above identification and had no previous history of knee joint injury. Participants were excluded if they had: (1) cognitive impairment with a score< 24 in the Mini-Mental State Examination, (2) involvement of the neurological diseases that affect walking ability, (3) involvement of severe dysfunction of the mean organs.

### Equipment

The spatial-temporal gait parameters and kinematic parameters in the paretic and the non-paretic legs were collected using VICON Motion Analysis System (Oxford Metrics Limited, UK) with eight infrared cameras and were analyzed by its supporting software system. The reflective markers were placed on the anatomical positions based on Plug in gait model (Figure 1). The sampling rate was 200Hz. Additionally, two Kistler force platforms were used to measure the ground reaction force and knee flexion/extension torques in the paretic and intact legs. The sampling rate was 1000 Hz. For measuring the water content of meniscus and the volume of meniscus, a GE Pioneer 3.0T Magnetic resonance imaging (MRI) scanner with a dedicated 16-channel knee surface coil and 3D dual echo UTE sequence was used for MRI examination for both subjects’ paretic and non-paretic knees. The scanning parameters were TR/TE1/TE2: 12/0/4.6 ms (respectively); Matrix: 400 × 400; field of view:180 mm; 2.0 mm thickness; Volume of voxel: 0.4 5x 0.45 × 2.0 mm3; Flap Angle: 8°; Half bandwidth:125 kHz. NEX=1. The positioning line was placed perpendicularly to the posterior edge of the femoral internal and external condyle, and 20 layers of sagittal images were collected consistently. The scanning time of each knee joint was 22 s.

**Figure 1.**
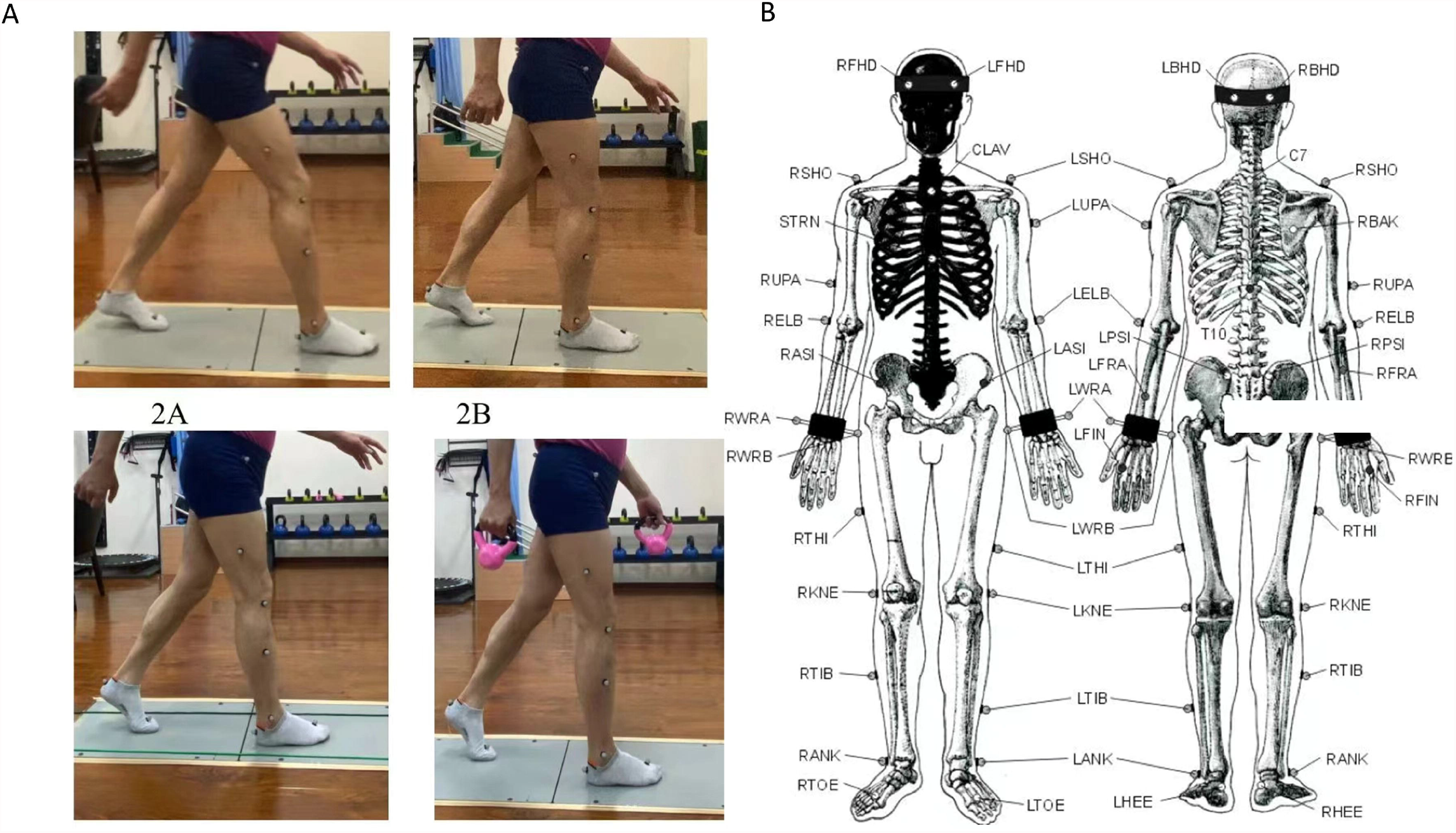
A) using a VICON Motion Analysis System to measure the gait, kinematic and kinetic parameters when a stroke patient walk through two force platforms; B) the marker setting for this study.

### Protocol

After participants were consented, they were sent to Magnetic resonance imaging room at Beijing Rehabilitation Hospital of Capital Medical University for imaging processing and data measurement. These images were captured and processed by radiologists with at least 10 years above experience in this field. To accurately measure the images, the steps needed to be followed. The first step was to open the TE = 0 images through the three-dimensional synchroview and select the optimal angle to measure the cartilage. Specifically, the signal intensity of cartilage at the thickest bearing point of the medial and lateral femoral condyles, the thickest bearing point of the medial and lateral tibial condyles, the anterior/posterior junction of the femur in the horn of the medial and lateral meniscus, and the anterior/posterior junction of the tibia in the horn of the medial and lateral meniscus were measured. Also, an elliptical region of interest was used to eliminate the errors for measuring the cartilage volume.

Then the second step was to open the TE = 4.6 ms images and repeated the above measurement process. The third step was to compare the images of TE = 0 and the images of TE = 4.6 ms. In the images of TE = 0, the signal intensity of cartilage was mainly formed by hydrogen protons of free water and bound water. In the TE = 4.6 ms images, the signal intensity of cartilage was mainly formed by hydrogen protons in free water. Thus, the percentage of free water content in deep and shallow layers could be calculated by the formula: I (TE = 4.6 ms) /I (TE = 0 ms) ×100%.

Each participant was instructed to walk through a 10-meter walkway with their comfortable speeds. At least two successful trials were used for data analyzing, indicating that participants needed to successfully step on each force platform with one leg only. If participant stepped on each force platform partially or stepped on one force platform with any portions of two legs, that specific trial will be noted as “Not Use”. Maximum attempted trials were setup at ten trials to prevent from fatigue in these patients. Between each trial, a two-minute mandatory rest was given to participants. Spatial-temporal parameters were quantified for both limbs (paretic and non-paretic): velocity, cadence, step length, step width, and stance phase duration. Kinematic parameters were quantified in both legs: peak knee extension; peak knee flexion; peak hip extension; peak hip flexion; peak ankle plantarflexion; and peak ankle dorsiflexion. Also, the maximum knee flexion/extension torques were quantified for both limbs. Finally, the average vertical ground reaction force was used for both limbs.

### Statistical analysis

A Pearson correlation was used to define the strength of the linear relationship between the angle of knee hypertension and one of abovementioned dependent variables. The coefficient of determination, R-squared, was used to understand the strength of the correlation. R-squared value < 0.09 indicated no or very weak relationship and R-squared value between 0.09 and 0.25 was weak relationship. R-squared value between 0.25 and 0.49 was moderate relationship. Finally, R-squared value larger than 0.49 indicated strong relationship. R value < 0.3 indicated no or very weak relationship and R value between 0.3 and 0.5 was weak relationship. R value between 0.5 and 0.7 was moderate relationship. Finally, R value larger than indicated strong relationship.

## RESULTS

### The correlation between angle of knee hyperextension and gait parameters, kinematics, kinetics

A moderate-to-strong correlation was found between the month and the angle of knee hyperextension (R = 0.56, *p* = 0.016), between the angle of knee hyperextension and the hip flexion in non-paretic leg (R = 0.42, *p* = 0.092), between the angle of knee hyperextension and the knee extension in paretic leg (R = 0.98, *p* < 0.001), between the angle of knee hyperextension and the ground reaction force in paretic leg (R = 0.58, *p* = 0.048), and between the angle of knee hyperextension and the knee torque in paretic leg (R = 0.48, *p* = 0.05). More detail was listed in the Table 1.

**Table 1.** The correlation between angles of hyperextension and gait parameters, kinematic or kinetic parameters.

| Gait Parameters |  | Kinematic |  | Kinetic |  |
| --- | --- | --- | --- | --- | --- |
|  | Hyperextension |  | Hyperextension |  | Hyperextension |
| Month | **R = 0.56, p = 0.016** | HipExtension Non-Paretic Leg | R = 0.15, p = 0.566 | Ground Reaction Force Non-Paretic Leg | R = 0.03, p = 0.930 |
| Step Length Non-Paretic Leg | R = 0.23, p = 0.351 | HipExtension Paretic Leg | R = 0.05, p = 0.843 | Ground Reaction Force Paretic Leg | **R = 0.58, p = 0.048** |
| Step Length Paretic Leg | R = 0.26, p = 0.301 | HipFlexion Non-Paretic Leg | **R = 0.42, p = 0.092** | Knee Torque Non-Paretic Leg | R = 0.22, p = 0.398 |
| Cadence Non-Paretic Leg | R = 0.03, p = 0.920 | HipFlexion Paretic Leg | R = 0.02, p = 0.930 | Knee Torque Paretic Leg | **R = 0.48, p = 0.050** |
| Cadence Paretic Leg | R = 0.07, p = 0.788 | KneeExtension Non-Paretic Leg | R = 0.20, p = 0.432 |  |  |
| Gait Speed Non-Paretic Leg | R = 0.20, p = 0.427 | KneeEXtension Paretic Leg | **R = 0.98, p < 0.001** |  |  |
| Gait Speed Paretic Leg | R = 0.23, p = 0.354 | KneeFlexion Non-Paretic Leg | R = 0.20, p = 0.434 |  |  |
| Stance Phase Non-Paretic Leg | R = 0.16, p = 0.951 | KneeFlexion Paretic Leg | R = 0.21, p = 0.431 |  |  |
| Stance Phase Paretic Leg | R = 0.28, p = 0.269 | AnkleDorsiFlexion Non-Paretic Leg | R = 0.24, p = 0.348 |  |  |
|  |  | AnkleDorsiFlexion Paretic Leg | R = 0.08, p = 0.767 |  |  |
|  |  | AnklePlantarFlexion Non-Paretic Leg | R = 0.08, p = 0.752 |  |  |
|  |  | AnklePlantarFlexion Paretic Leg | R = 0.29, p = 0.253 |  |  |

### The correlation between knee hyperextension and meniscus volume

A moderate-to-strong correlation was found between the angle of knee hyperextension and the medial meniscus volume in non-paretic leg (R = - 0.53), between the angle of knee hypertension and the medial meniscus volume in paretic leg (R = - 0.57), and between the angle of knee hypertension and the lateral meniscus volume in paretic leg (R = - 0.70). More detail was shown in Figure 2 and Figure 3.

**Figure 2.**
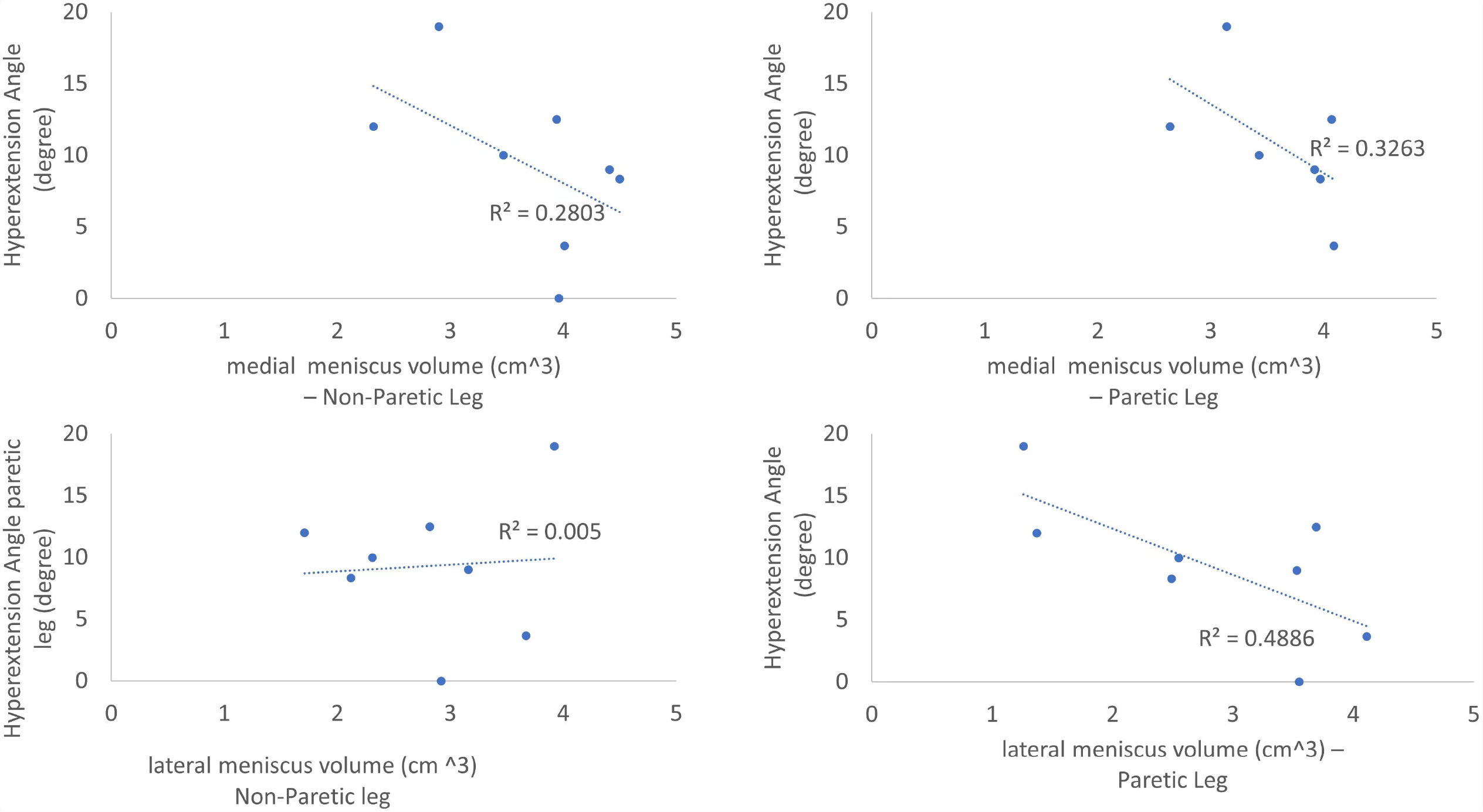
The correlations between angles of knee hyperextension and meniscus volume.

**Figure 3.**
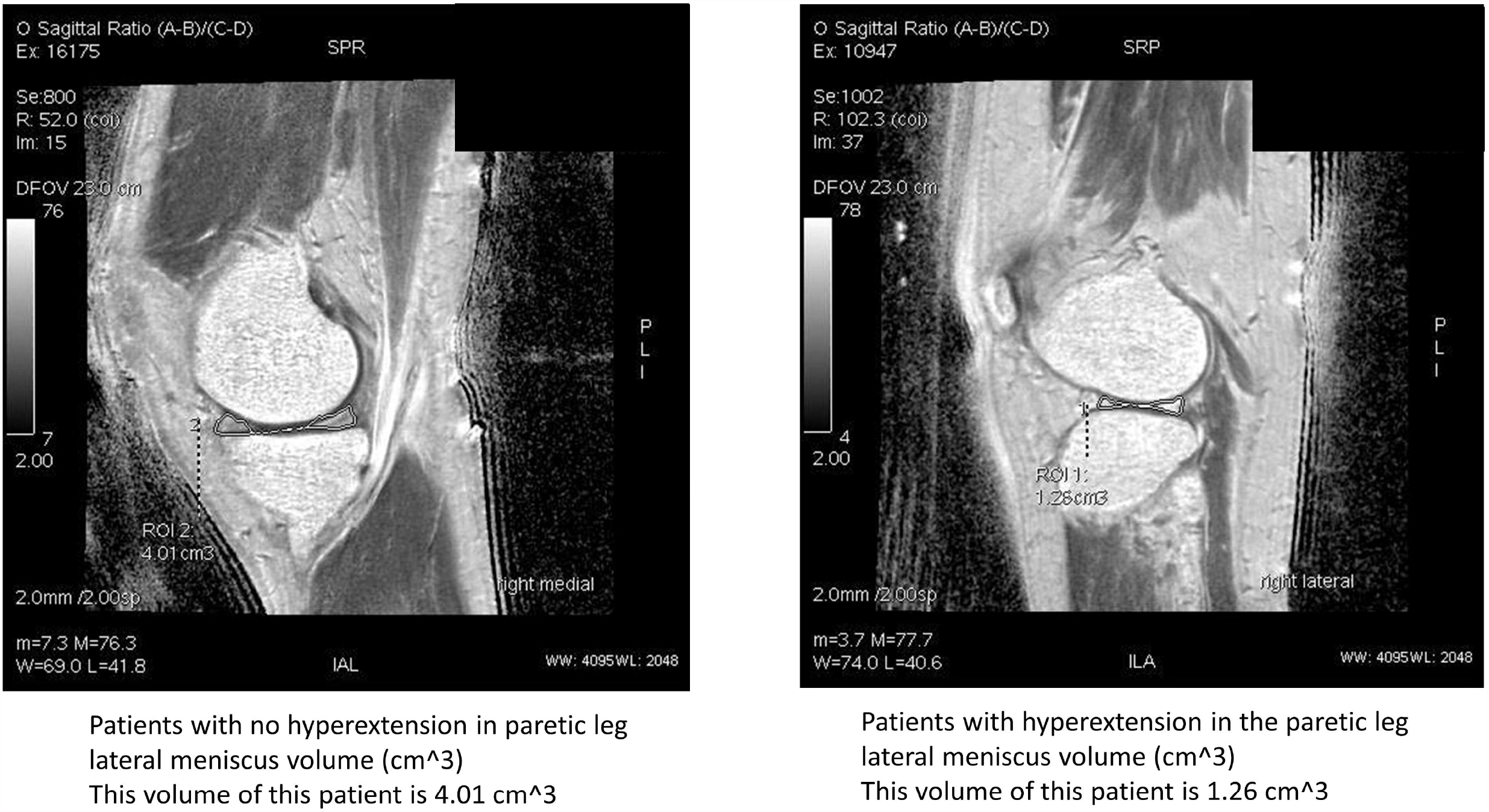
The meniscus volume between two stroke patients: no hyperextension in paretic leg (Left side) and had a hyperextension in paretic leg (Right side).

### The correlation between knee hyperextension and water content of meniscus

A moderate-to-strong correlation was found between the angle of knee hyperextension and the water content of anterior angle of lateral meniscus in non-paretic leg (R = -0.91), between the angle of knee hyperextension and the water content of anterior angle of medial meniscus in paretic leg (R = 0.53), and between the angle of knee hyperextension and, the water content of posterior angle of medial meniscus in paretic leg (R = 0.63). More detail was shown in Figure 4.

**Figure 4.**
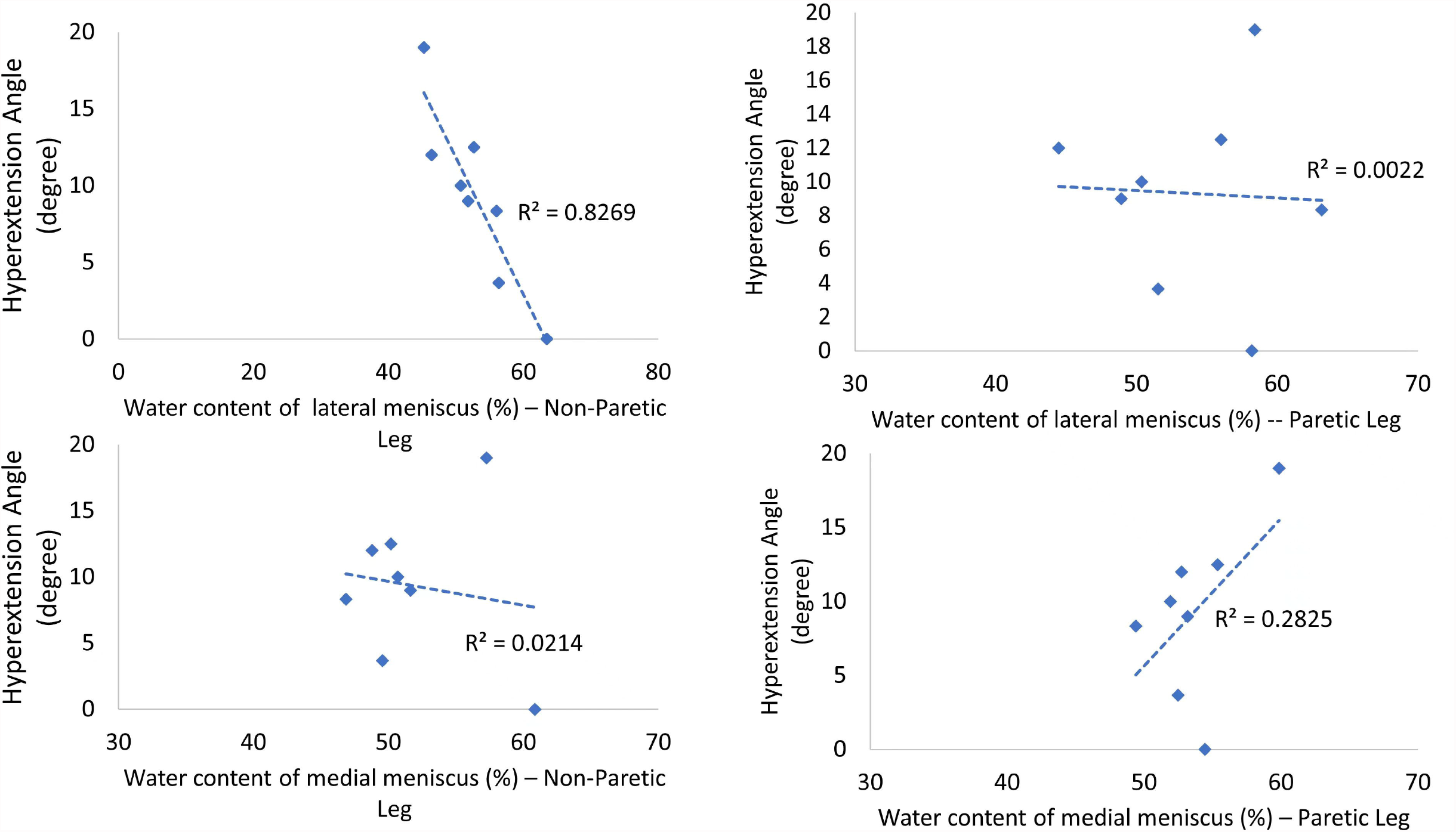
The correlations between angles of knee hyperextension and water content of meniscus.

## DISCUSSION

To our best knowledge, this study was the first study to investigate the corrections between the angles of hyperextension and the gait parameters, joint angles, kinetic parameters. Also, the correlations between the angles of knee hypertension and meniscus volume, and the water content of meniscus were investigated. Surprisingly, the results in this study found low correlations between the angles of knee hypertension and gait parameters, joint angles, and kinetic parameters. However, the results found that higher knee hyperextension indeed had a strong-to-moderate negative correlations between the angles of hyperextension and medial/lateral meniscus volume in paretic legs.

### Longer time since hemiplegia led to larger angles of knee hyperextension

It has been reported that approximate half of stroke survivors experienced the knee hyperextension. Importantly, this knee hyperextension may be caused by muscle weakness and the impairment of proprioceptive sensors [11]; furthermore, this knee hyperextension causes pain and limit the patients’ movement in daily life activities [8]. However, in past decades, it has been widely investigated the symptoms of the knee hyperextension in stroke survivors. For instance, this knee hyperextension generates the knee extensor moment to prevent from falls during stance phase. However, it has never been shown the correlation between the time since hemiplegia and the angles of knee hyperextension in the previous literature. In the current study, a positive correlation was observed between the time since hemiplegia and the angles of knee hyperextension. In other words, longer time since suffering hemiplegia, larger angles of knee hyperextension could be gained. Therefore, it is worth mentioning this correlation because the best treatment exist for knee hypertension in stroke patients at the current moment is the Ankle-foot orthoses. However, a study also shows that in patients with severe knee hyperextension, the Ankle-Foot orthoses might not be effective [13]. Using Knee-Ankle-Foot orthosis might an appropriate alternative [13]. However, the drawbacks of the Knee-Ankle-Foot could delay the recovery of normal gait and increase the spasticity of the gait in stroke survivors [13]. In such a case, based on the observation in this study, to identify the time since hemiplegia can be an initial screen to identify whether the Ankle-Foot or Knee-Ankle-Foot orthoses.

### Larger angles of knee hyperextension led to more tears in medial/lateral meniscus

The medial and lateral menisci anatomically cover the superior aspect of tibia. The lateral meniscus is more circular than the medial meniscus; however, the medial meniscus is more crescent-like than the lateral meniscus. The roles of menisci are to bear the stress across the knee during standing or walking. Importantly, these menisci are to prevent knee hyperextension during walking. However, when putting too much pressure on the knee joint, the knee joint is forced to extend further than the healthy range of motion resulting in potential tears of the ligaments in stroke patients. This might be the rationales that a negative correlation between the angles of hyperextension and the meniscus volume was found in both paretic legs in the current study. Specifically, due to the shape of medial meniscus, the medial meniscus in general suffers three times more stress than the lateral meniscus during walking [12]. In the current study, a negative correlation between the angles of hyperextension and the medial meniscus volume was observed, indicating that higher hyperextension might lead to more tears in the medial meniscus not only in the paretic leg but also in the non-paretic leg. This result might raise a concern that the knee hyperextension might need to be seriously treated as soon as possible.

### Larger angles of knee hyperextension decreased water content of the lateral meniscus in the non-paretic leg but increased water content of the medial meniscus

Firstly, the average water contents were 52.85%, 53.89%, 51.98% and 53.68% of the lateral meniscus – Non-Paretic, lateral meniscus – Paretic Leg, medial meniscus – Non-Paretic, and medial meniscus – Paretic Leg, respectively. These mean values were much lower than normal healthy controls (72%) [14]. It was not a surprising result, indicating no matter knee hyperextension existed or not, the level of water content apparently decreased by the meniscus degradation due to the stroke [15]. The interesting results were that larger angles of knee hyperextension had a tendency to decrease water content of the lateral meniscus than in the non-paretic leg but had a tendency to increase water content of the medial meniscus in the paretic leg. This could be explained by Warnecke et al., study that as increasing degrees of degenerations of medial meniscus in the paretic leg, the water content significantly increases and further leads to worsen viscoelastic properties inside the knee [16]. This phenomenon might reduce the water content of lateral meniscus in the non-paretic leg due to the compensated mechanism.

### The angles of knee hyperextension and gait parameters had a no or low correlation

The alternations of gait parameters due to the quadriceps weakness, quadriceps spasticity, calf muscles spasticity have been linked to the knee hyperextension. Surprisingly, in the current study, a low or no correlation between angles of knee hypertension and step length, cadence, or walking speed. Similar results have found in a review with 44 relevant studies, involving 2658 patients with stroke, concludes that stroke survivors who receive treadmill are not likely to improve their ability in their step length, cadence, and stance phase [17]. However, using joint angles and kinetic parameters might be better options to identify the knee hyperextension than spatial-temporal gait parameters. Importantly, in the current study, larger knee hyperextension in the paretic leg induced larger hip flexion in non-paretic leg but not the angle joint to provide a compensation for balance. This result inferred that for stroke patients, the leading joint was the proximal joint because these proximal joints could generate larger power to maintain balance and also moved the body forward.

### Clinical implication

The clinical implications for this study were 1) the knee hyperextension needed to be diagnosed and treated as early as possible; 2) the time since hemiplegia could be an indicator of sign of knee hyperextension; and 3) it might be not effective to identify the knee hyperextension by using spatial-temporal gait parameters.

### Limitation

The apparent limitation was the sample size. Currently, only eight stroke survivors were included in this study. In the future, more stroke survivors need to be recruited to identify the results in this study. However, this study was the first using MRI to demonstrate larger knee hyperextension led to more tears in medial/lateral meniscus and larger water content of meniscus.

## Data Availability

All files are available from the database of Beijing Rehabilitation Hospital

## Authors’ contributions

Wenshan Li and Tiancong Li wrote the main text. Wenshan Li and Tiancong Li designed the experiment and Wenshan Li, Tiancong Li, Xiaoshuang Xi and Rong Zhang carried out the experiment. Weishuang Sun, Conglin Han and Dan Zhang processed and analyzed the data. All authors reviewed and agreed the content of this manuscript.

